# Sex disparities in authorship in pulmonary hypertension research: a scoping review protocol

**DOI:** 10.1101/2021.11.21.21266658

**Authors:** Kathryn Lalonde, Shannon Ruzycki, Lisa Mielniczuk, Jason Weatherald

## Abstract

From protocol introduction: “Our aim is to examine female authorship and sex disparities in the domain of pulmonary hypertension (PH) research. Despite PH disproportionately affecting females, we hypothesize that the proportion of studies with women as first or senior author will be < 50% and women will be underrepresented in publications in more prestigious journals.”

## INTRODUCTION

In many aspects of research and medicine there has been increasing attention on sex and gender equality, opportunity and achievement. Over time, there has been an increasing number of females being admitted into medical school and subsequently more female physicians in many countries, including Canada, the United States and Britain^1,2,3^. However, the increasing proportion of female physicians does not necessarily equate to equal leadership opportunity or authorship in academia^4,5,6,7,8^. Less favorable assessments of women as principal investigators may explain lower success in grant funding^9^. These biases in academia may lead to inequitable opportunities for and inadequate representation of female across different roles.

Several recent articles examined gender and sex disparities in the pulmonary literature. One paper by Kawano-Dourado et al. (2020) examined female representation in Interstitial Lung Disease (ILD) research which underlined that female Academics are under-represented in leadership roles, that is likely perpetuated by explicit and implicit bias^10^. Another study of interest by Vranas et al. (2020) examined the sex difference in authorship in the critical care literature^11^. This paper highlighted that far fewer women were publishing and those that did were more likely to publish as co-authors and/or in lower impact journals as compared to their male counterparts. There was also very little increase in the proportion of female first or senior authors between 2009-2018.

Our aim is to examine female authorship and sex disparities in the domain of pulmonary hypertension (PH) research. Despite PH disproportionately affecting females, we hypothesize that the proportion of studies with women as first or senior author will be < 50% and women will be underrepresented in publications in more prestigious journals.

### Objectives

1. To describe the proportion of manuscripts with a female first or senior author in the PH peer-reviewed literature.
2. To describe the proportion of female first or senior-authored PH manuscripts among articles in the top 20 journals searched, according to journal impact factor.
3. To determine the proportion of manuscripts with a female first or senior author among the top 500 most cited papers in PH.
4. To describe temporal trends in female authorship in the PH literature.

## METHODS

### Study Design

This study will be a scoping review of the literature that will follow the PRISMA-ScR reporting recommendations^12^. This protocol will be published on a pre-print server prior to initiation of article screening.

### Search Strategy

We will perform a broad search in Medline/OVID. The search terms will include:

- “Pulmonary Hypertension” in title or abstract (with no adjacency)
- “Pulmonary Arterial Hypertension” in title or abstract (with no adjacency)

We have chosen this strategy as it allows capture of other important disease synonyms, including “Primary pulmonary hypertension” and related diseases such as “Chronic Thromboembolic Pulmonary Hypertension”. We will pilot our search results by ensuring 17 landmark PH papers with female first or senior authorship (**Appendix 1**) are captured in the search results.

### Inclusion Criteria

#### 1) Publication date between January 1, 2000 – November 20, 2021

When examining the density of published literature across time, there are many more articles published in PH in more recent years. For that reason, we have decided to include a large time span of 20 years as it will enable us to evaluate trends over a longer time interval without markedly increasing the workload to an unmanageable level.

#### 2) Journal title

We will restrict articles to those published in the 80 pre-specified journals (**Appendix 2**), in order to make the number of citations manageable. We have purposively selected a range of journals, including respiratory medicine and cardiology journals, surgery, general medical, basic science, and pediatric journals. These journals have varying degrees of number of articles published on pulmonary hypertension and a wide range of impact factors. This list of journals is based on a consensus between two content experts (LM and JW). We will ensure that each journal has at least 1 article present after the data set has been refined by date (no journals with 0 publications on PH will be included).

#### 3) Adult or Pediatric PH population

We have included 9 pediatric journals to specifically account for the pediatric population.

#### 4) Article type

We will limit inclusion to articles that are clinical trials (including randomized control trials), observational studies, basic science/translational reports, systematic reviews with or without meta-analysis, guidelines, narrative review articles, case report/series, and editorials.

### Exclusion Criteria

#### 1) Commentaries, Replies/Responses

These will not be included as they are often opinion-based rather than original research.

#### 2) Books and Book Chapters

These will not be included as they often have solitary authors contributing to each chapter; the importance order of authorship is not always clear when there are multiple authors listed.

#### 3) Conference Presentations and Poster Abstracts

Though this would provide insight into who may be participating in current PH research, it would not answer our question of women’s authorship in peer review publications for PH.

#### 4) Language Restriction

We will not apply a language filter. Non-English articles will have title and abstract translated with Google Translate to screen suitability. For those articles unable to be translated sufficiently, these will be excluded from the overall data set.

### Screening Process

We will screen articles by title and abstract for relevance by one team member. Given the large number of anticipated search results, screening for relevance will proceed after applying inclusion criteria filters (**Figure 1**).

**Figure 1.**
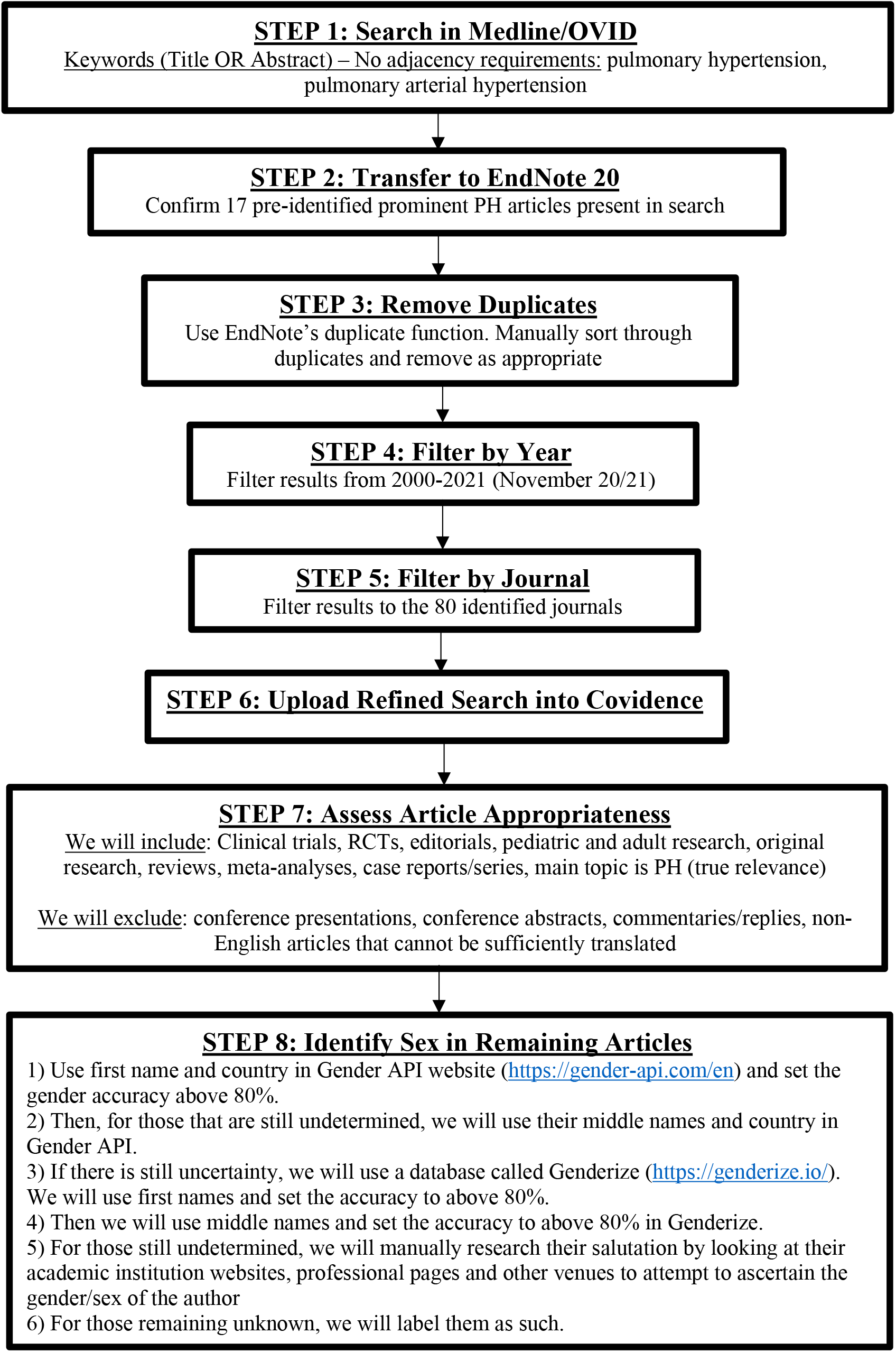
Study Selection process.

## Data Management

The search results from Medline/OVID will be imported into EndNote 20. In EndNote, the articles will be screened for duplicates. Exact duplicates will be removed. If articles have been published in more than one journal, they will be included. Replies and editorials will be kept if identified as duplicates at this stage. Then the article data will be filtered by year – including 2000-2021 (November 1). Then, the selected journals will be identified and filtered as such.

The paired down search results will be imported to Covidence.org, where duplicates will be identified and removed (as a secondary check). Then, this data set will be sorted through by Dr. Lalonde and Dr. Weatherald, where they will each follow the pre-set inclusion and exclusion criteria. As this is not a review paper, we will not require that each article is screened twice (i.e., each article will be screened once).

The final data will be exported from Covidence into Excel for analysis.

### Data Extraction

We will extract the following information from each article that meet the eligibility criteria.

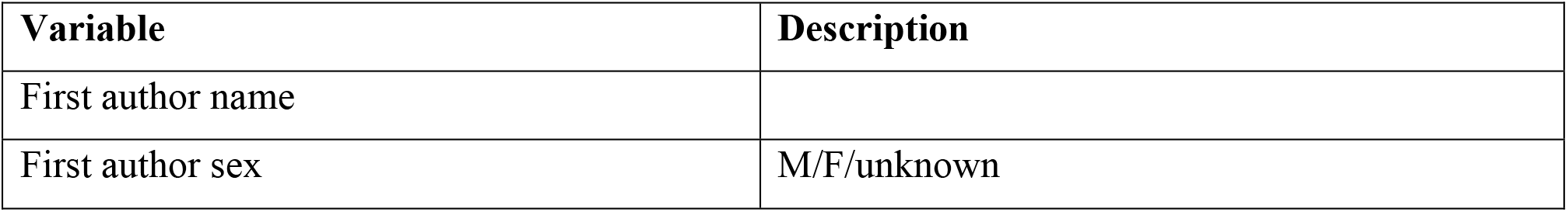
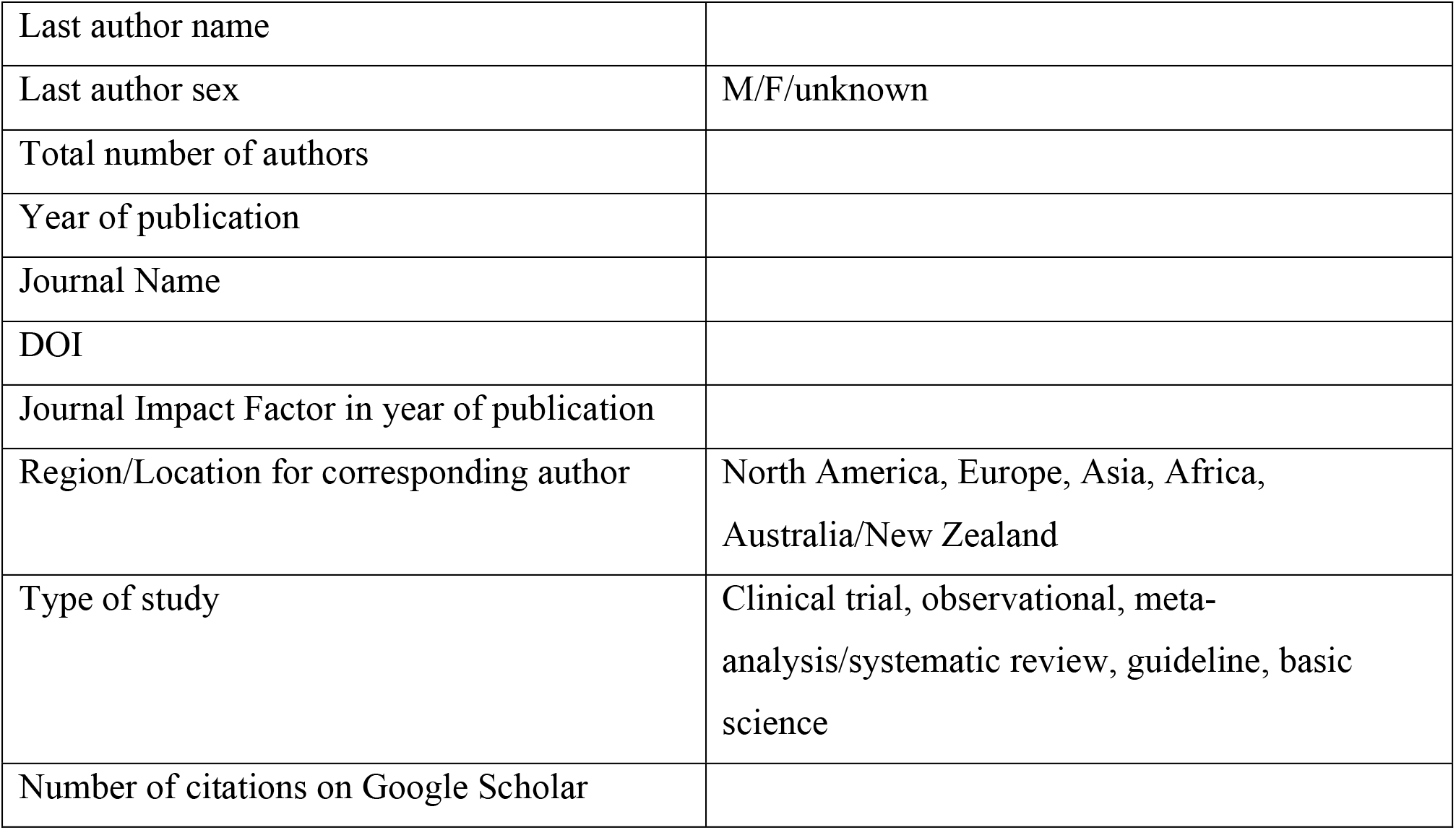

| **Variable** | **Description** |
| --- | --- |
| First author name |  |
| First author sex | M/F/unknown |
| Last author name |  |
| Last author sex | M/F/unknown |
| Total number of authors |  |
| Year of publication |  |
| Journal Name |  |
| DOI |  |
| Journal Impact Factor in year of publication |  |
| Region/Location for corresponding author | North America, Europe, Asia, Africa,<br>Australia/New Zealand |
| Type of study | Clinical trial, observational, meta-analysis/systematic review, guideline, basic<br>science |
| Number of citations on Google Scholar |  |

### Sex Determination

A study by Gayet-Ageron *et al* (2021) evaluated female authorship in the COVID-19 literature^13^. We will use similar methodology in our sex determination strategy:

1. Use first name and country in *Gender API* website (https://gender-api.com/en) and set the gender accuracy above 80%.
2. Then, for those that are still undetermined, we will use their middle names and country in Gender API.
3. If there is still uncertainty, we will use a database called *Genderize* (https://genderize.io/). We will use first names and set the accuracy to above 80%.
4. Then we will use middle names and set the accuracy to above 80% in *Genderize*.
5. For those still undetermined, we will manually research their salutation by looking at their academic institution websites, professional pages and other venues to attempt to ascertain the sex of the author
6. For those remaining unknown, we will label them “unknown”.

*Genderize* was also used in the study by Vranas et al. (2020)^11^. When the sex of authors is uncertain we will attempt to access academic institution websites, professional pages and other online articles to ascertain the sex of the authors, where possible.

### Outcomes and Analysis

The primary outcome is the proportion of all articles with female first or senior authorship. Secondary outcomes include:

1. Proportion of articles with female first authorship
2. Proportion of articles with female senior/last authorship
3. Proportion of articles with female authorship per year (2000-2021)
4. Proportion of articles with female first or senior authorship across quartiles or quintiles of journal impact factors.
5. Proportion of articles with female first or senior authorship across the top 30 cited papers in PH per year.
6. Number of contributors on female first/senior authored papers compared to male first/senior authored papers
7. Female first/senior authorship across different subtypes of studies (RCTs vs. Observational, Clinical vs. Basic Science, etc).

We will assess the association between sex of first/senior authorship (dependent variable) with journals impact factor and continent of publication using logistic regression. We will assess the association between female senior authorship and sex of the first author using logistic regression. We will stratify analyses by clinical and basic science manuscripts.

Given the international nature of scientific research and lack of gendered names in same languages or countries, it is possible that we may not be able to identify the sex of all authors. We will do a sensitivity analysis excluding articles with unknown sex of either first or last author, or with “anonymous” authorship, or with authors limited to working groups/steering committee titles.

## Data Availability

This is currently just a protocol of a study to be conducted. All data produced in the present study will be available upon reasonable request to the authors.

## Funding

There is no funding to report for this study.

## Author Contributions

KL and JW conceived this study.

KL, JW, SR, and LM designed the methods.

KL and JW wrote the study protocol.

Final study protocol has been approved by all authors.

### Appendix 2 – List of Journal Titles for Inclusion

#### Adult Medicine Focus

1. Journal of the American College of Cardiology
2. Circulation
3. JAMA Cardiology
4. European Heart Journal
5. European Journal of Heart Failure
6. JACC: Heart Failure
7. Circulation Research
8. Nature Reviews Cardiology
9. Circulation: Heart Failure
10. American Heart Journal
11. American Journal of Cardiology
12. Hypertension
13. Cardiovascular Research
14. Chest
15. Journal of the American Heart Association
16. Heart
17. International Journal of Cardiology
18. Canadian Journal of Cardiology
19. American Journal of Cardiovascular Disease
20. Lancet – Respiratory Medicine
21. American Journal of Respiratory and Critical Care Medicine
22. Journal of Heart and Lung Transplantation
23. European Respiratory Journal
24. Thorax
25. European Respiratory Review
26. Annals of the American Thoracic Society
27. Respirology
28. Respiratory Research
29. Respiratory Medicine
30. BMC Pulmonary Medicine
31. Pulmonary Circulation
32. Respiration
33. Intensive Care Medicine
34. Critical Care Medicine
35. Critical Care
36. Journal of Critical Care
37. Journal of Intensive Care
38. Annals of Intensive Care
39. JAMA Internal Medicine
40. Annals of Internal Medicine
41. Journal of Internal Medicine
42. Journal of General Internal Medicine
43. New England Journal of Medicine
44. Lancet
45. The BMJ
46. Nature
47. Nature Communications
48. Nature Medicine
49. Nature Clinical Practice Cardiovascular Medicine
50. Nature Clinical Practice Rheumatology
51. Nature Genetics
52. Nature Reviews Cardiology
53. Nature Reviews Molecular Cell Biology
54. Nature Reviews Rheumatology
55. Nature Reviews Drug Discovery
56. Nature Structural and Molecular Biology
57. American Journal of Physiology – Heart and Circulatory Physiology
58. American Journal of Physiology – Lung Cellular and Molecular Physiology
59. Journal of Applied Physiology
60. Journal of Clinical Investigation
61. Annals of the Rheumatic Diseases
62. Arthritis & Rheumatology
63. Rheumatology
64. JACC: Cardiovascular Imaging
65. Journal of the American Society of Echocardiography
66. Circulation: Cardiovascular Imaging
67. European Heart Journal Cardiovascular Imaging
68. BMJ Open Respiratory Research
69. Pulmonary Pharmacology and Therapeutics
70. Journal of Thoracic and Cardiovascular Surgery
71. Cochrane Database of Systematic Reviews

#### Pediatric Medicine Focus

1. JAMA Pediatrics
2. Pediatrics
3. Journal of Pediatrics
4. Frontiers of Pediatrics
5. Pediatric Research
6. BMC Pediatrics
7. Pediatric Pulmonology
8. Pediatric Cardiology
9. Pediatric Critical Care Medicine

## Notes

### Competing Interest Statement

The authors have declared no competing interest.

### Funding Statement

This protocol did not receive any funding.

